# How many calories are in our food? Compliance with England’s calorie labelling regulations three years after policy implementation

**DOI:** 10.1101/2025.07.03.25330811

**Authors:** Oliver Huse, Alexandra Kalbus

## Abstract

**Objective:** To assess compliance with England’s calorie labelling regulations in the out-of-home (OOH) food sector.

**Methods:** Data were collected from websites of large UK OOH food outlets. Descriptive analysis of compliance with the calorie labelling regulations was conducted, particularly i) whether the ‘default menu’, i.e. the first menu a consumer is likely to see, shows calories, ii) whether implementation guidance relating to label visibility and statement of daily calorie needs were followed, and iii) whether compliance varied by business type.

**Results:** While all outlets (n=77) provided calorie labelling somewhere on their websites, just under half (48%) did not provide calorie labels on the default menu, requiring additional steps to view a labelled menu. Compliance with the policy’s guidance was greatest for the label’s position (81%) and lowest for prominent formatting (40%), while 71% provided the statement of daily calorie needs. Differences between OOH outlet types were observed but not tested owing to the small sample size.

**Conclusions:** Our results suggest that OOH food outlets are finding ways to circumvent the calorie labelling regulations, thus undermining the policy’s impact. As the policy’s review is approaching, policymakers should consider strategies for ensuring compliance.

**Plain Language Title and Summary:** *Assessing how well food businesses in England are following Calorie labelling laws requiring them to show Calorie information on menus:* To address the rising prevalence of diet-related disease, the English government introduced mandatory calorie labelling on the menus of restaurants and other large food outlets. This study looked at how well food outlets in England are following the rules about showing calorie information on their menus. Researchers checked the websites of 77 large food outlets in the UK to see if they were adhering to the calorie labelling laws. They focused on three main things: 1. Whether the first menu a customer sees (the “default menu”) clearly shows calorie information.
2. Whether the calorie labels are easy to see and include a reminder about how many calories a typical adult needs in a day.
3. Whether certain types of food businesses are adhering to the law better than others. Although all 77 businesses did include calorie information somewhere on their websites, nearly half of them (48%) did not show calories on the first menu customers would see. Instead, customers had to click through extra steps to find the calorie-labelled menu, which could discourage them from using it. Most businesses (81%) put calorie information in the correct location on the menu, but only 40% made it clearly visible with easy-to-read formatting. About 71% of businesses correctly included the reminder that adults need around 2,000 calories per day. Different types of food businesses were following the rules to varying degrees, but the sample size was too small to draw firm conclusions. Overall, the study suggests that many food businesses are not fully following the calorie labelling laws, and some may be doing the minimum to technically comply while making it harder for customers to access calorie information. As the government reviews this policy, better enforcement mechanisms are needed to ensure that businesses comply with the regulations.

## Introduction

Diets high in energy-dense and nutrient-poor foods are a key contributor to poor health globally (1). In the UK, food prepared away from home contributes on average 300 kilocalories (kcal) per person per day (2). Meals served in large UK restaurant and fast-food chains are particularly calorie dense, reaching almost 1000 kcal per serving on average (3).

To help consumers make informed choices when purchasing food prepared out of home (OOH), mandatory calorie labelling was introduced in England on 6^th^ April 2022 (4). All businesses with ≥250 employees are required to display the calorie content of food and non-alcoholic drink items prepared for immediate consumption at the point of choice (including online) (4). The calorie label must correspond to the serving size offered, be displayed in kcal and accompanied by the statement ‘adults need around 2,000 kcal a day’, referred to as statement of daily calorie needs (4).

Evidence suggests that the calorie labelling regulations have had minimal impact on consumer purchasing (5, 6). One possible reason for this is low compliance by OOH food outlets – most businesses only partially comply with government regulations (7), and enforcement mechanisms are lacking (8). Just one study (7) has assessed compliance with the guidelines, considering only in-store retail a few months post policy implementation. The aim of this study was to assess compliance with calorie labelling regulations among large OOH food outlets in England three years post policy implementation, and to assess whether compliance varies by OOH food outlet type.

## Methods

OOH food chains investigated were aligned with those investigated by MenuTracker, a longitudinal database of online menus (9). OOH food chains were included if their website included a menu which consumers might view to make a food purchasing decision. OOH food chains that were aimed at business-to-business retail, or that did not present a food menu online, were not included.

Websites for all included OOH food chains were visited by one of two authors (OH or AK) between 1^st^ and 4^th^ April 2025, from the same location. Both authors collected data for a subset of 20 websites to assess inter-rater reliability. Websites were visited in private browser mode and the online food menu was accessed via the most direct pathway to assess compliance with the calorie labelling regulations. If a website required a location, a standardised postcode (WC1H 9SH) was provided, and the closest available outlet was selected.

Descriptive data analysis was carried out in R version 4.5.0. Supplementary Material 1 and Supplementary Material 2 contain the data collected and code used in this study, respectively.

## Results

In total, 77 OOH food chains were included in this study: 16 ‘cafes and bakeries’, 20 ‘pubs, bars and inns’, 17 ‘restaurants’, 23 ‘fast-food and takeaway outlets’, and 1 ‘sport and entertainment venue’. The latter was excluded where data were stratified by outlet type. Among the included OOH food chains, all displayed calorie labelling on at least one of the menus available on their website. Just over half (52%, n=40) of OOH food chains showed calorie labelling on their default menu (Figure 1). Of those that did not (n=37), 57% (n=21) required customers to click on the menu item, 32% required labelled menus to be downloaded either through a click (n=11) or a QR code (n=1), and 11% (n=4) required customers to choose a branch-specific menu or start an online order.

**Figure 1.**
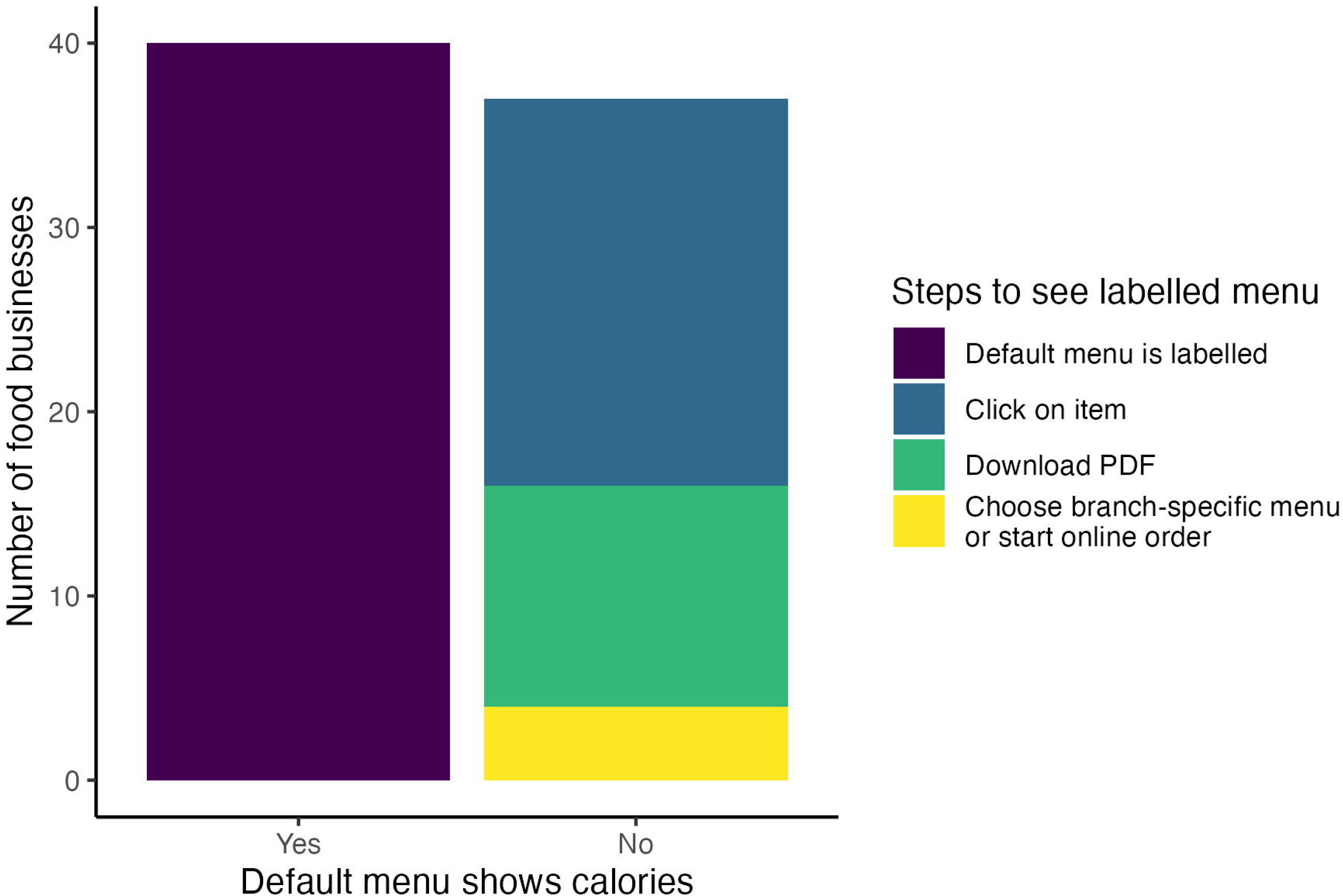

Calorie labels were displayed for all eligible items by 79% (n=61) of OOH food outlets (Figure 2). Compliance was greatest in terms of the calorie label’s position (close to the item’s name, price or description) with 81% (n=62) of OOH food outlets. However, just 40% (n=31) of food outlets presented calorie labelling in at least as prominent font and size as the item name, price or description. The statement of daily calorie needs was present and correct on the menus of 71% (n=55) of assessed OOH food outlets. Overall, 20% (n=15) of businesses were compliant with all assessed criteria, of which only one was a fast-food outlet. Observed differences by food outlet type were not tested for statistical significance due to the small sample size.

**Figure 2.**
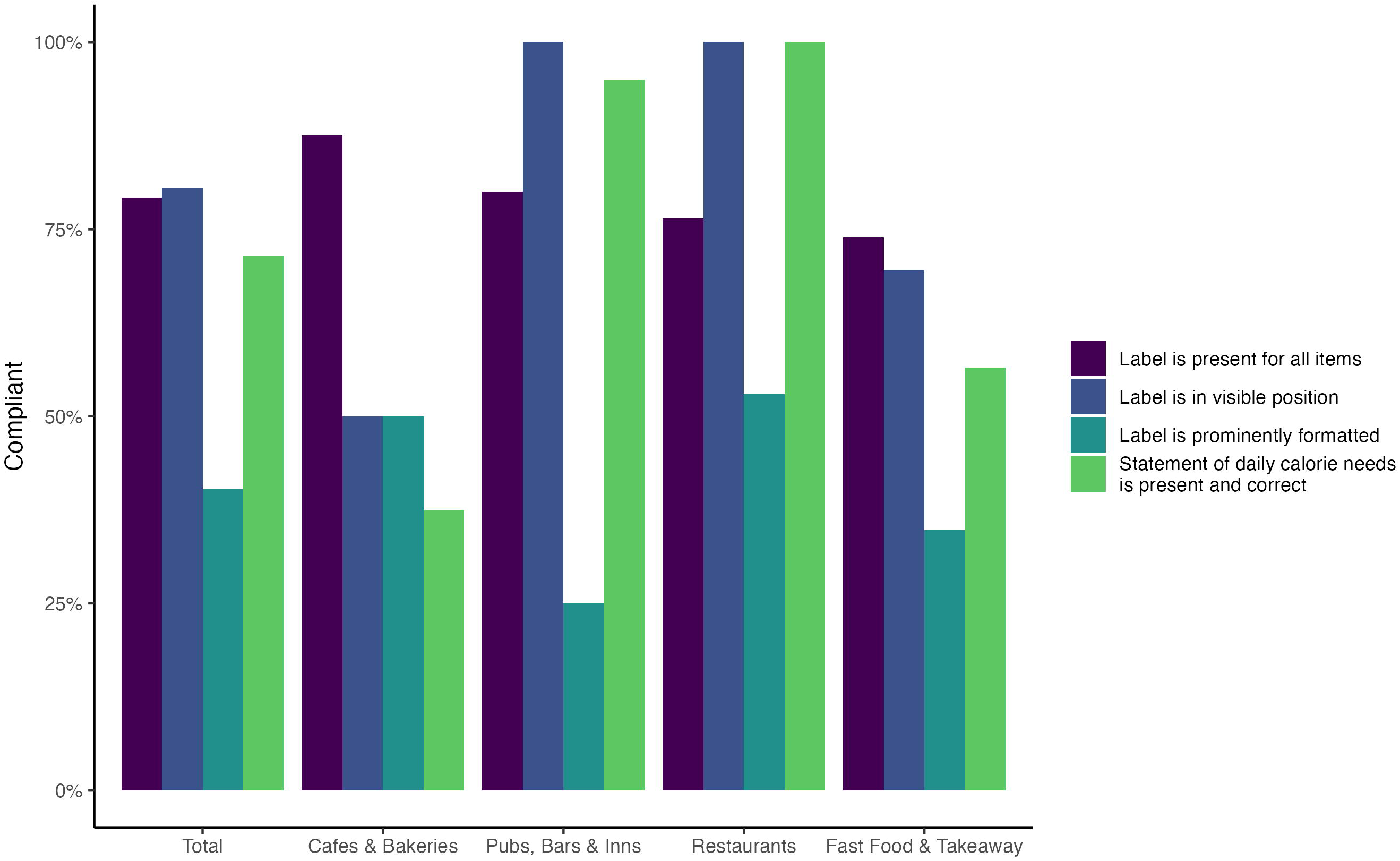

## Discussion

Whilst all studied OOH food outlets provided calorie information on their websites, many employed tactics to avoid displaying calories to the consumer at the point of choice. Just under half of the included OOH food businesses did not display calorie labelling on the ‘default’ menu, i.e. the first point of choice, but instead requiring additional steps to view calorie-labelled menus. Calorie labels were frequently displayed in a less prominent font style, size or colouring compared to other menu text. Likewise, several used alternative wordings for the statement of daily calorie needs or did not display this at all.

Previously, Polden et al. (7) reported that a few months after policy implementation, 80% and 67% of outlets provided calorie labelling on any and all menus available in-store, respectively, while only 6% provided prominently formatted calorie labelling, and 45% provided a clear and prominent statement of daily calorie needs (7). Our study builds upon this work by assessing the menus consumers encounter online three years post policy implementation and also considering the ‘default’ menu. Considering the reported lack of enforcement capacity from local government (8), it is not surprising that OOH food outlets are not consistently complying with the calorie labelling regulations.

To the best of our knowledge, this is one of just two studies to assess compliance with England’s calorie labelling regulations. However, several limitations apply. While unlikely, assessed online menus may not represent in-store menus. We only included businesses listed in the MenuTracker database (9) and cannot comment on other large businesses. Finally, this cross-sectional analysis cannot assess shifts in compliance over time.

Our results suggest that OOH food outlets are finding ways to circumvent the calorie labelling regulations, thus undermining the policy’s impact. As the policy’s review is approaching (4), it will be important for the UK government to consider i) how to ensure compliance, and ii) other strategies, including alternative labelling options, fiscal approaches, and marketing restrictions, to best support consumers in achieving a healthy diet in OOH food settings.

## Data Availability

All data produced in the present study are contained in the supplementary materials of this manuscript.

## Acknowledgments

The authors wish to thank Dr Yuru Huang and Dr Michael Essman for their advice on the MenuTracker database, from which businesses included in this report were sampled.

## Notes

### Competing Interest Statement

The authors have declared no competing interest.

### Funding Statement

This study did not receive any funding.

