## Supplementary Material 2 for "How many calories are in our food? Compliance with England’s calorie labelling regulations three years after policy implementation"

```
############# Calorie labelling compliance analysis ################

# load packages
library(dplyr) # data manipulation (incl %>% pipe)
library(ggplot2) # for plots
library(here) # relative folder location - no need to type full file paths if script in same folder as data/other outputs
library(readxl) # read in excel files
library(viridis) # for colour palette
library(scales) # for the percent stacked bar chart (to print % on y-axis)

# load data
d <- read_excel(here("Calorie labelling compliance data.xlsx")) # we can use shorter file path here relative to where the the script is saved

# View data
View(d)

## Data prep ####
# drop 'notes' row and variables not needed in this analysis, and shorten variable names
d <- d %>%
  slice(-1) %>% # removes the first row (notes)
  select(`Restaurant name`, Type, `Labelling provided anywhere`, `Default menu shows calories`, `Steps to see labelled menu`, `Labelled menu more than 1 click away`, `Labelled menu more than 2 clicks away`, `Labels present for all items`, `Label position`, `Label prominence`, `Reference statement`, `Labels shown when ordering online`) %>%
  rename(`Statement of daily calorie needs` = `Reference statement`)

count(d) # 77 outlets
table(d$`Labelling provided anywhere`)

# we grouped Western & Asian Fast Food (n=3) together
# for stratification by business type, we excluded the 1 sport 7 entertainment venue (n=76)
d_type <- d %>%
  filter(Type != "Sport & Entertainment") %>%
  mutate(Type = recode(Type,
                       "Asian Fast Food" = "Fast Food & Takeaway",
                       "Cafes and Bakeries" = "Cafes & Bakeries",
                       "Pubs, Bars & Inns" = "Pubs, Bars & Inns",
                       "Restaurants" = "Restaurants",
                       "Western Fast Food & Takeaway" = "Fast Food & Takeaway"))

## Summary stats ####
d_type %>% group_by(Type) %>% count() # plus 1 sport & entertainment venue

### Does the default menu display calories? ####
d %>%
  group_by(`Default menu shows calories`) %>%
  summarise(`Number of restaurants` = n()) %>%
  mutate(`%` = `Number of restaurants` / sum(`Number of restaurants`) * 100)

# by restaurant type type
d_type %>%
  group_by(`Type`, `Default menu shows calories`) %>%
  summarise(`Number of restaurants` = n()) %>%
  mutate(`% default menu shows kcal` = `Number of restaurants` / sum(`Number of restaurants`) * 100) %>%
  filter(`Default menu shows calories` == 'Yes') %>%
  select(Type, `% default menu shows kcal`)

# chi-square test by business type (not reported due to small sample size)
#contingency_table <- table(d_type$Type, d_type$`Default menu shows calories`)
#contingency_table # same table as above - chi square command needs a table as input
#chisq.test(contingency_table)

### Where the default does not show calories, what are the necessary steps to see calorie labels (n=37)? ####
d %>%
  filter(`Default menu shows calories`=='No') %>%
  group_by(`Steps to see labelled menu`) %>%
  summarise(`Number of restaurants` = n()) %>%
  mutate(`%` = `Number of restaurants` / sum(`Number of restaurants`) * 100)

# by business type
d_type %>%
  filter(`Default menu shows calories`=='No') %>%
  group_by(`Type`,`Steps to see labelled menu`) %>%
  summarise(`Number of restaurants` = n()) %>%
  mutate(`%` = `Number of restaurants` / sum(`Number of restaurants`) * 100)

### How many clicks to get to labelled menu where this is not the default? ####
# Is the labelled menu more than 1 click away?
d %>%
  filter(`Default menu shows calories`=='No') %>%
  group_by(`Labelled menu more than 1 click away`) %>%
  summarise(`Number of restaurants` = n()) %>%
  mutate(`%` = `Number of restaurants` / sum(`Number of restaurants`) * 100)

# Is the labelled menu more than 2 clicks away?
d %>%
  filter(`Default menu shows calories`=='No') %>%
  group_by(`Labelled menu more than 2 clicks away`) %>%
  summarise(`Number of restaurants` = n()) %>%
  mutate(`%` = `Number of restaurants` / sum(`Number of restaurants`) * 100)

### Is labelling available for all eligble items? ####
d %>%
  group_by(`Labels present for all items`) %>%
  summarise(`Number of restaurants` = n()) %>%
  mutate(`%` = `Number of restaurants` / sum(`Number of restaurants`) * 100)

# by business type
d_type %>%
  group_by(`Type`, `Labels present for all items`) %>%
  summarise(`Number of restaurants` = n()) %>%
  mutate(`% compliant` = `Number of restaurants` / sum(`Number of restaurants`) * 100) %>%
  filter(`Labels present for all items` == 'Yes') %>%
  select(Type, `% compliant`)

# save % for compliance graph
c_items <- d_type %>%
  group_by(`Type`, `Labels present for all items`) %>%
  summarise(`Number of restaurants` = n()) %>%
  mutate(`%` = `Number of restaurants` / sum(`Number of restaurants`) * 100) %>%
  filter(`Labels present for all items` == 'Yes') %>%
  select(Type, `%`) %>%
  mutate(compliance = "Label is present for all items")
all_c <- d %>%
  group_by(`Labels present for all items`) %>%
  summarise(`Number of restaurants` = n()) %>%
  mutate(`%` = `Number of restaurants` / sum(`Number of restaurants`) * 100,
         Type = "Total",
         compliance = "Label is present for all items") %>%
  filter(`Labels present for all items` == 'Yes') %>%
  select(Type, `%`, compliance)

### How visible is the calorie label (position)? ####
d %>%
  group_by(`Label position`) %>%
  summarise(`Number of restaurants` = n()) %>%
  mutate(`%` = `Number of restaurants` / sum(`Number of restaurants`) * 100)

# by business type
d_type %>%
  group_by(`Type`, `Label position`) %>%
  summarise(`Number of restaurants` = n()) %>%
  mutate(`% compliant` = `Number of restaurants` / sum(`Number of restaurants`) * 100) %>%
  filter(`Label position` == 'Close to name, price or description') %>%
  select(Type, `% compliant`)

# save % for compliance graph
c_position <- d_type %>%
  group_by(`Type`, `Label position`) %>%
  summarise(`Number of restaurants` = n()) %>%
  mutate(`%` = `Number of restaurants` / sum(`Number of restaurants`) * 100) %>%
  filter(`Label position` == 'Close to name, price or description') %>%
  select(Type, `%`) %>%
  mutate(compliance = "Label is in visible position")
all_c <- d %>%
  group_by(`Label position`) %>%
  summarise(`Number of restaurants` = n()) %>%
  mutate(`%` = `Number of restaurants` / sum(`Number of restaurants`) * 100,
         Type = "Total",
         compliance = "Label is in visible position") %>%
  filter(`Label position` == 'Close to name, price or description') %>%
  select(Type, `%`, compliance) %>% rbind(all_c)

### How prominently formatted is the label? ####
d %>%
  group_by(`Label prominence`) %>%
  summarise(`Number of restaurants` = n()) %>%
  mutate(`% compliant` = `Number of restaurants` / sum(`Number of restaurants`) * 100)

# by business type
d_type %>%
  group_by(`Type`, `Label prominence`) %>%
  summarise(`Number of restaurants` = n()) %>%
  mutate(`% compliant` = `Number of restaurants` / sum(`Number of restaurants`) * 100) %>%
  filter(`Label prominence` == 'Yes') %>%
  select(Type, `% compliant`)

# save % for compliance graph
c_prominence <- d_type %>%
  group_by(`Type`, `Label prominence`) %>%
  summarise(`Number of restaurants` = n()) %>%
  mutate(`%` = `Number of restaurants` / sum(`Number of restaurants`) * 100) %>%
  filter(`Label prominence` == 'Yes') %>%
  select(Type, `%`) %>%
  mutate(compliance = "Label is prominently formatted")
all_c <- d %>%
  group_by(`Label prominence`) %>%
  summarise(`Number of restaurants` = n()) %>%
  mutate(`%` = `Number of restaurants` / sum(`Number of restaurants`) * 100,
         Type = "Total",
         compliance = "Label is prominently formatted") %>%
  filter(`Label prominence` == 'Yes') %>%
  select(Type, `%`, compliance) %>% rbind(all_c)

### Statement of daily calorie needs ####
d %>%
  mutate(`Statement of daily calorie needs compliant` = ifelse(`Statement of daily calorie needs`=="Statement is present and clear" | `Statement of daily calorie needs`=="Statement is in fineprint", "Yes", "No")) %>%
  group_by(`Statement of daily calorie needs compliant`) %>%
  summarise(`Number of restaurants` = n()) %>%
  mutate(`%` = `Number of restaurants` / sum(`Number of restaurants`) * 100)

# by business type
d_type %>%
  mutate(reference_compliant = ifelse(`Statement of daily calorie needs`=="Statement is present and clear" | `Statement of daily calorie needs`=="Statement is in fineprint", "Yes", "No")) %>%
  group_by(`Type`, reference_compliant) %>%
  summarise(`Number of restaurants` = n()) %>%
  mutate(`% compliant` = `Number of restaurants` / sum(`Number of restaurants`) * 100) %>%
  filter(reference_compliant == 'Yes') %>%
  select(Type, `% compliant`)

# save % for compliance graph
c_reference <- d_type %>%
  mutate(reference_compliant = ifelse(`Statement of daily calorie needs`=="Statement is present and clear" | `Statement of daily calorie needs`=="Statement is in fineprint", "Statement of daily calorie needs", "not compliant")) %>%
  group_by(`Type`, reference_compliant) %>%
  summarise(`Number of restaurants` = n()) %>%
  mutate(`%` = `Number of restaurants` / sum(`Number of restaurants`) * 100) %>%
  filter(reference_compliant == "Statement of daily calorie needs") %>%
  select(Type, `%`) %>%
  mutate(compliance = "Statement of daily calorie needs \nis present and correct")
all_c <- d %>%
  mutate(reference_compliant = ifelse(`Statement of daily calorie needs`=="Statement is present and clear" | `Statement of daily calorie needs`=="Statement is in fineprint", "Statement of daily calorie needs", "not compliant")) %>%
  group_by(reference_compliant) %>%
  summarise(`Number of restaurants` = n()) %>%
  mutate(`%` = `Number of restaurants` / sum(`Number of restaurants`) * 100,
         Type = "Total",
         compliance = "Statement of daily calorie needs \nis present and correct") %>%
  filter(reference_compliant == "Statement of daily calorie needs") %>%
  select(Type, `%`, compliance) %>% rbind(all_c)

### Compliance with all criteria ####
# default menu + all items labelled + label formatting & position + statement
d %>%
  mutate(reference_compliant = ifelse(`Statement of daily calorie needs`=="Statement is present and clear" | `Statement of daily calorie needs`=="Statement is in fineprint", "Statement of daily calorie needs", "not compliant"),
         `Compliant with all criteria` = ifelse(`Default menu shows calories` == "Yes" & `Labels present for all items` == "Yes" & `Label position` == "Close to name, price or description" & `Label prominence`== "Yes" & reference_compliant == "Statement of daily calorie needs", "Yes", "No")) %>%
  group_by(`Compliant with all criteria`) %>%
  summarise(`Number of restaurants` = n()) %>%
  mutate(`% compliant with all criteria` = `Number of restaurants` / sum(`Number of restaurants`) * 100) %>%
  filter(`Compliant with all criteria` == 'Yes') %>%
  select(`Number of restaurants`,`% compliant with all criteria`)

# By business type
d_type <- d_type %>%
  mutate(reference_compliant = ifelse(`Statement of daily calorie needs`=="Statement is present and clear" | `Statement of daily calorie needs`=="Statement is in fineprint", "Statement of daily calorie needs", "not compliant"),
         `Compliant with all criteria` = ifelse(`Default menu shows calories` == "Yes" & `Labels present for all items` == "Yes" & `Label position` == "Close to name, price or description" & `Label prominence`== "Yes" & reference_compliant == "Statement of daily calorie needs", "Yes", "No"))

d_type %>%
  group_by(Type,`Compliant with all criteria`) %>%
  summarise(`Number of restaurants` = n()) %>%
  mutate(`% compliant with all criteria` = `Number of restaurants` / sum(`Number of restaurants`) * 100) %>%
  filter(`Compliant with all criteria` == 'Yes') %>%
  select(Type,`Number of restaurants`,`% compliant with all criteria`)

# chi-square test by business type (not reported due to small sample size)
#contingency_table <- table(d_type$Type, d_type$`Compliant with all criteria`)
#contingency_table # same table as above - chi square command needs a table as input
#chisq.test(contingency_table)

### Ordering online ####
d %>%
  group_by(`Labels shown when ordering online`) %>%
  summarise(`Number of restaurants` = n()) %>%
  mutate(`%` = `Number of restaurants` / sum(`Number of restaurants`) * 100)

# by business type
d_type %>%
  group_by(`Type`, `Labels shown when ordering online`) %>%
  summarise(`Number of restaurants` = n()) %>%
  mutate(`%` = `Number of restaurants` / sum(`Number of restaurants`) * 100)

## Graphs #####
### default menu with labelling & steps to get to it ####

# create data for plotting
d_graph <- d %>%
  group_by(`Default menu shows calories`, `Steps to see labelled menu`) %>%
  summarise(value = n()) %>%
  mutate(`Default menu shows calories` = factor(`Default menu shows calories`, levels = c("Yes", "No")), # makes the 'yes' bar appear first
         `Steps to see labelled menu` = factor(ifelse(is.na(`Steps to see labelled menu`), "Default menu is labelled", recode(`Steps to see labelled menu`,
                                               "Click on item" = "Click on item",
                                               "choose branch-specific menu or start online order" = "Choose branch-specific menu \nor start online order",
                                               "download pdf" = "Download PDF",
                                               "download pdf - with QR code" = "Download PDF")),
                                               levels = c("Default menu is labelled", "Click on item", "Download PDF", "Choose branch-specific menu \nor start online order")))

# View the underlying numbers
d_graph

colours = viridis(4) # set graph colours
ggplot(d_graph, aes(fill=`Steps to see labelled menu`, y=value, x=`Default menu shows calories`)) + # tell R we want to make a figure (ggplot) with which data and variables to use
  geom_bar(position="stack", stat="identity") + # specify what type of graph
  scale_fill_manual(values = colours) + # specify colours (of 'fill' - here the steps to see the labelled menu); taken from the viridis palette
  labs(y = "Number of food businesses") + # specify axis title
  theme_classic() # set general formatting
#ggsave(here("Fig 1. Steps to see labelled menu.tiff"), width = 6, height=4) # saves the last produced figure into the specified file, in specified format and dimensions

### default menu showing calories by business type (not shown in report) ####
total_graph <- d %>%
  mutate(Type = "Total") %>%
  group_by(Type, `Default menu shows calories`) %>%
  summarise(value = n()) %>%
  mutate(pct= prop.table(value) * 100)
d_graph <- d_type %>%
  group_by(Type, `Default menu shows calories`) %>%
  summarise(value = n()) %>%
  mutate(pct= prop.table(value) * 100) %>%
  rbind(total_graph) %>%
  mutate(Type = factor(Type, levels = c("Total", "Cafes & Bakeries", "Pubs, Bars & Inns", "Restaurants", "Fast Food & Takeaway")))

ggplot(d_graph, aes(x=Type, y=value, fill=`Default menu shows calories`)) +
  geom_bar(position="fill", stat="identity") +
  scale_fill_manual(values = c("#31688EFF", "#35B779FF")) +
  labs(y = NULL, x = NULL) +
  theme_classic() +
  scale_y_continuous(labels = scales::percent)
#ggsave(here("Type and default menu percent stacked.tiff"), width = 8.5, height = 5)

### Labelling compliance graph ####
d_graph <- rbind(c_items, c_position, c_prominence, c_reference, all_c) %>% # row-binds datasets created above
  mutate(Type = factor(Type, levels = c("Total", "Cafes & Bakeries", "Pubs, Bars & Inns", "Restaurants", "Fast Food & Takeaway")),
         compliance = factor(compliance, levels = c("Label is present for all items", "Label is in visible position", "Label is prominently formatted", "Statement of daily calorie needs \nis present and correct"))) # the \n in the text makes the text break to the next line in figure

ggplot(d_graph, aes(x=Type, y=`%`/100, fill=compliance)) +
  geom_bar(position="dodge", stat="identity") +
  scale_fill_manual(values = viridis(5)) +
  labs(y = "Compliant", x = NULL, fill = NULL) + # fill=NULL removes legend title
  theme_classic() +
  scale_y_continuous(labels = scales::percent)
#ggsave(here("Fig.2 Compliance characteristics by type.tiff"), width=9, height=5.5)
```
